# The Role of Social Media in Disseminating Plastic Surgery Research: The Relationship between Citations, Altmetrics and Article Characteristics

**DOI:** 10.1101/2020.08.26.20182337

**Authors:** Michael C Grant, Kai R Scott-Bridge, Ryckie G Wade

## Abstract

**Background:** Social media (SoMe) enables publishers and authors to disseminate content immediately and directly to interested end-users, on a global scale. Alternative metrics (altmetrics) are non-traditional bibliometrics which describe the exposure and impact of an article on freely available platforms such as Twitter, Facebook, Wikipedia and the news. Altmetrics are strongly associated with ultimate citation counts in various medical disciplines, except plastic surgery which represents the rational for this study.

**Methods:** Altmetric explorer was used to extract altmetrics and citation rates for articles published during 2018 in Plastic and Reconstructive Surgery (PRS), the Journal of Plastic, Reconstructive and Aesthetic Surgery, the Annals of Plastics Surgery and Plastic Surgery (also known as Chirurgie Plastique). Multivariable negative binomial regression was used to estimate the relationship between citations and predictors (presented as the incidence rate ratio, IRR with 95% confidence interval, CI). Results: Overall, 1215 plastic surgery articles were captured which were cited 3269 times. There was a strong and independent association between the number of mentions in SoMe and the number of times an article was cited (adjusted IRR 1.01 [95% CI 1.01, 1.1]), whereby each mention in SoMe (e.g. Tweets or Facebook posts) translated to one additional citation. Evidence synthesis articles (e.g. systematic reviews) were cited twice as often as other articles and again, the use of SoMe to advertise these outputs was independently associated with more citations (IRR 2.0 [95% CI 1.3, 3.2]).

**Conclusions:** Dissemination of plastic surgery research through social media channels increases an articles impact as measured by citations.

## Introduction

Social media (SoMe) offers a powerful alternative to the traditional routes of research article distribution. Normally, publishers actively disseminate journal articles directly to paid subscribers via printed or digital issues, as well as passively listing them online. However, the SoMe revolution and its numerous platforms (such as Twitter, Facebook, Instagram, etc) allow both publishers and authors to disseminate content immediately and directly to interested end-users on a global scale^1^. Moreover, SoMe platforms facilitate concise and contemporaneous discussion in the same thread, which might otherwise be relegated to the correspondence section of unrelated journal issues.

Twitter is the most popular microblogging site for medical research^2^ and global events^3^. Indeed, recent work has shown that plastic surgeons use Twitter primarily for professional purposes^4^. The character limit of Tweets (<280) encourages brevity and the use of hashtags (#) helps to filters traffic to personally relevant content^5^. With Twitter’s ever increasing usership, its sphere of influence over medial research continues to grow^6^.

The Impact Factor and SCImago Journal Rank are journal-level metrics used to indirectly measure the journals’ success in disseminating its content. The h-index is an author-level metric which quantifies the impact of an individual. The IF, SJR and h-index scores are all based upon citations because the more valuable an article, the more often it is cited in subsequent publications. Alternative metrics (altmetrics) are non-traditional bibliometrics which offer complementary information about the exposure and impact of an article beyond citations. Altmetrics quantify activities such as Tweets, Facebook and Instagram posts, reads on Mendeley, media coverage, mentions on medical blogs, citations in Wikipedia and much more. Therefore, altmetrics are increasingly valued by academics, institutions and journals7 because they represent both short and long-term attention for an article.

This study aims to explore 1) the impact of SoMe activity on citations in the short term, and 2) the SoMe activity associated with plastic surgery publications.

## Methods

Prominent plastic surgery journals were selected for this cross-sectional study, including: *Plastic and Reconstructive Surgery* (PRS; published by Lippincott Williams and Wilkins of Wolters Kluwer), the *Journal of Plastic, Reconstructive and Aesthetic Surgery* (JPRAS; published by Elsevier), *Annals of Plastics Surgery* (APS; published by Lippincott Williams and Wilkins of Wolters Kluwer) and *Plastic Surgery* (PS, also known as Chirurgie Plastique published by Sage). We selected PRS, JPRAS and APS as they were regularly active on SoMe by January 2018 whilst PS served as a comparator with no formal SoMe presence. Therefore, we sampled articles published in 2018.

During January 2020, the digital object identifier (DOI) of each article was input to the Altmetric Explorer (www.altmetric.com) and altmetrics data were extracted. If no data were available on Altmetric Explorer, then Scopus data was extracted. Articles were categorised by MG/KSG into: correspondence (letters to the Editor or short manuscripts lacking an abstract which were listed in the correspondence section of the journal), case reports, primary research (full length original articles with an associated abstract) or evidence synthesis articles (review article, whether performed systematically or non-systematically, with or without a resultant meta-analysis). Articles were also categorised by topic (subspecialty) by RGW. The Altmetric score is derived from an automated algorithm and represents a weighted count of the amount of attention an article received (Supplementary Table 1).

Data were analysed using Stata v15. Proportions were compared using the chi squared or Fisher exact test, as appropriate. Count data were widely dispersed so negative binomial regression was used to estimate the relationship between citation counts and predictors. The effect of co-variables on the rate of citations is presented as incident rate-ratios (IRR) with 95% confidence intervals (CI).

## Results

Overall, 1215 articles were captured during 2018 from 4 journals including: PRS (n=565), JPRAS (n=358), APS (n=258) and PS (n=34). These articles were cited 3269 times; 2149 citations (66%) were of articles published in PRS, 725 (22%) in JPRAS, 365 (11%) in APS and 30 (1%) in PS. PRS, JPRAS and APS had active Twitter accounts with 15,900, 1742 and 143 followers, respectively. The same three journals had Facebook accounts with 29,862, 711 and 270 likes respectively. PRS used two other SoMe platforms and JPRAS used one other platform.

The characteristics of articles published by each journal differed substantially (Table 1). Whilst all journals published more original research than any other article type, there were significant differences in the prevalence of open access mansucripts and the types of articles. There were no meaningful discrepancies in the prevalence of articles per topic.

**Table 1.**
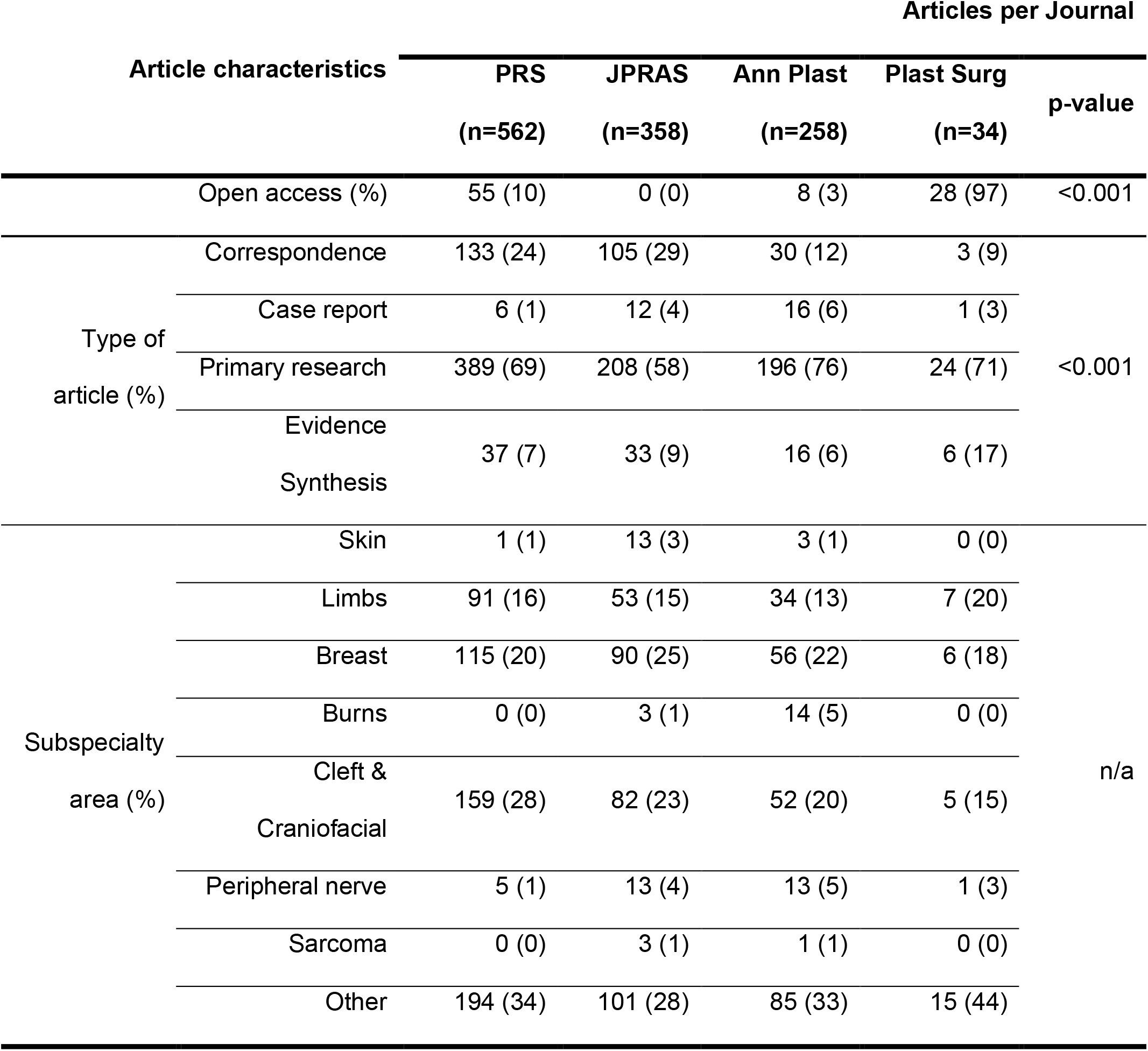
Journal publishing activity

Table 2 shows the citation counts and altmetrics for each journal. On average, articles published in PRS had at least one more citation (median difference 1 [IQR 1, 1], p<0.001) and an Altmetric score which was 3-points higher (median difference 3 [IQR CI 2, 5], p<0.001) than articles published elsewhere. On average, articles published in August, November and December had half the number of citations than articles published during the other months of the year (IRR 0.54 [95% CI 0.45, 0.64]; Supplementary Figure 1).

**Table 2.**
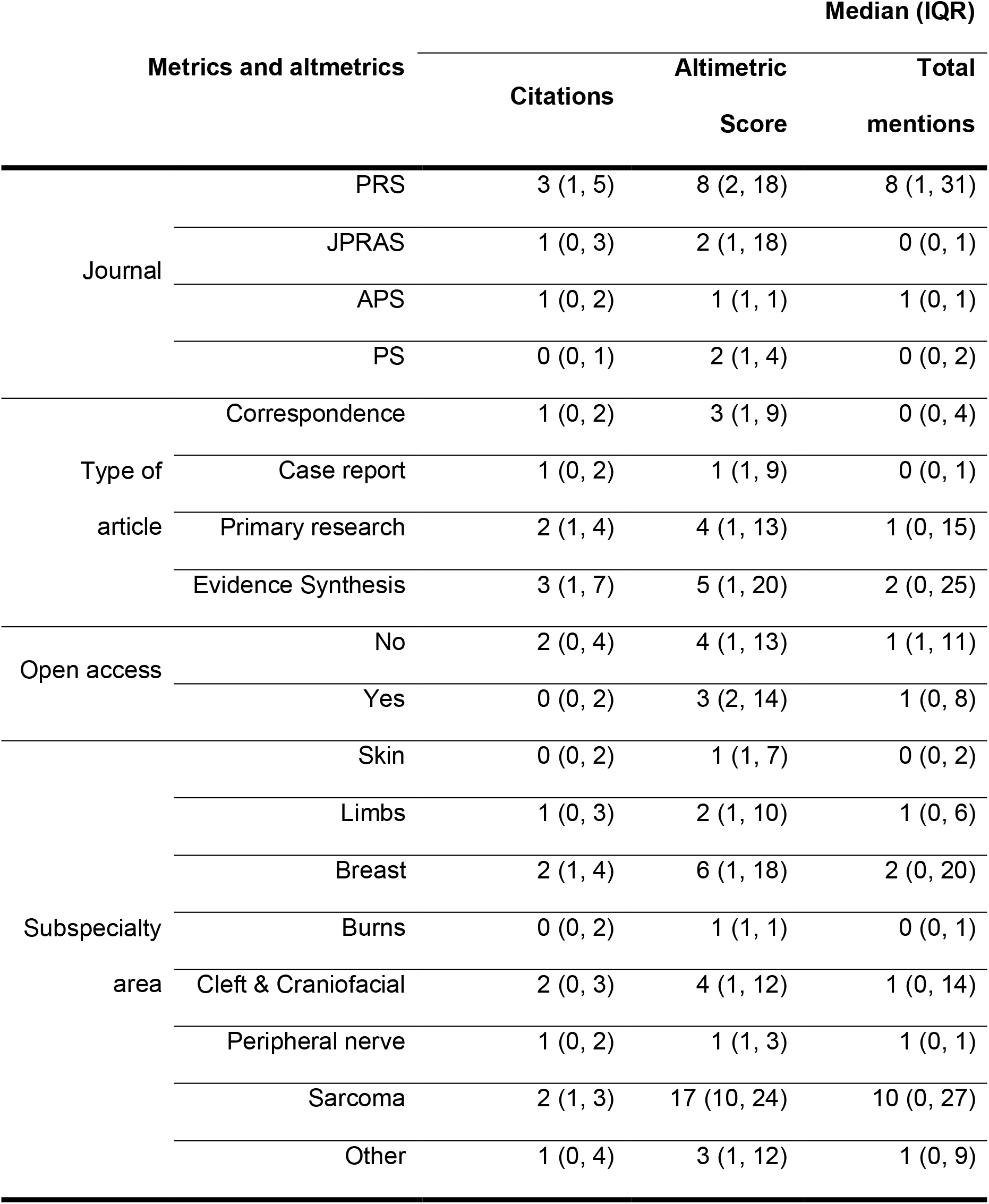
Metrics and altmetrics

Multivariable analysis (Table 3) showed that the use of SoMe to disseminate research was independently associated with more citations (Figure 1), whereby every mention in SoMe translated to one additional citation. The journal in which articles were published was also strongly and independently related to citation counts (Figure 2). As expected, evidence synthesis articles (systematic reviews with or without meta-analysis) were cited approximately three-times as often as correspondence or case reports (IRR 2.9) and more often than any other type of article (Figure 3), with SoMe mentions independently associated with increased citation counts.

**Table 3.**
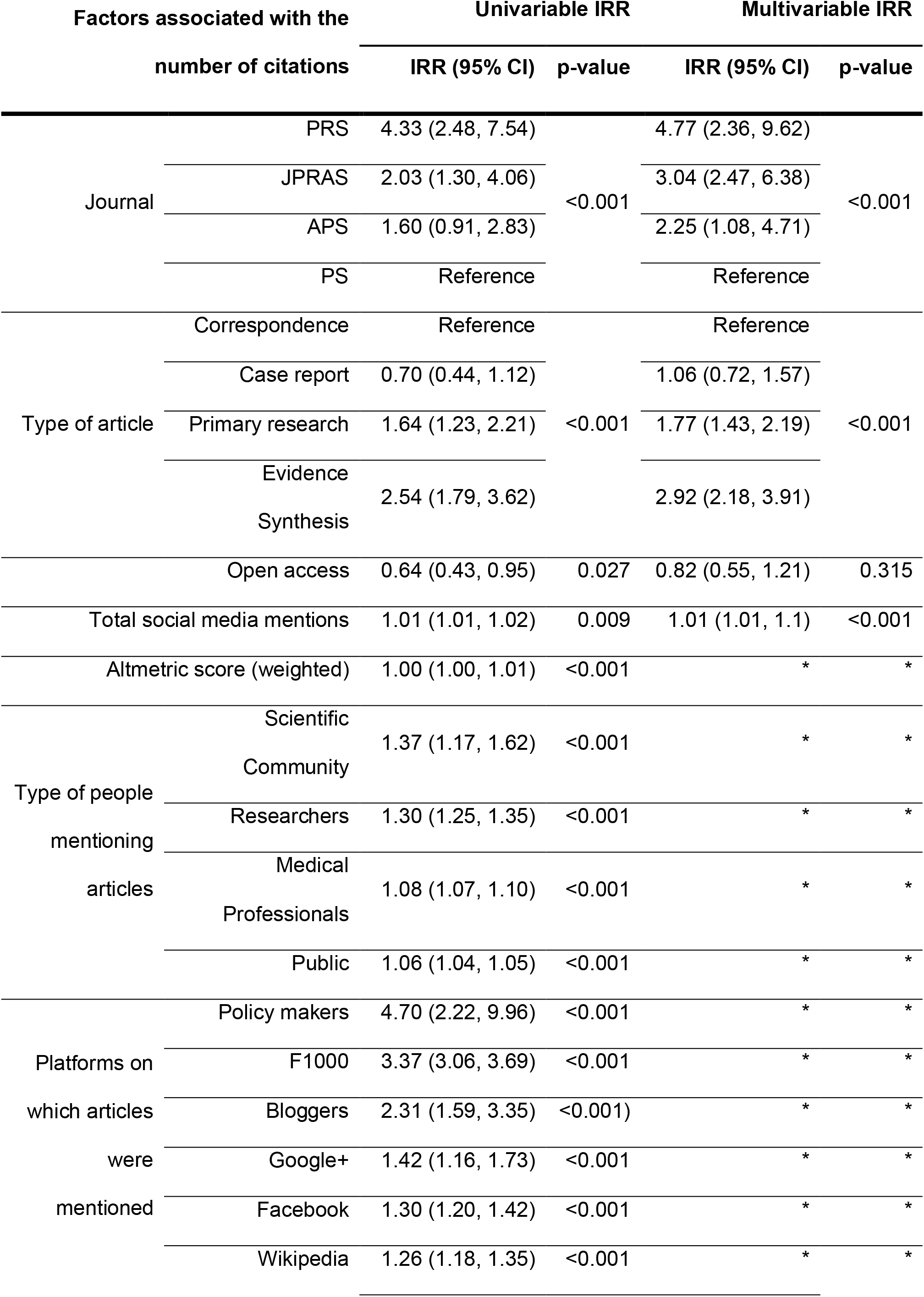
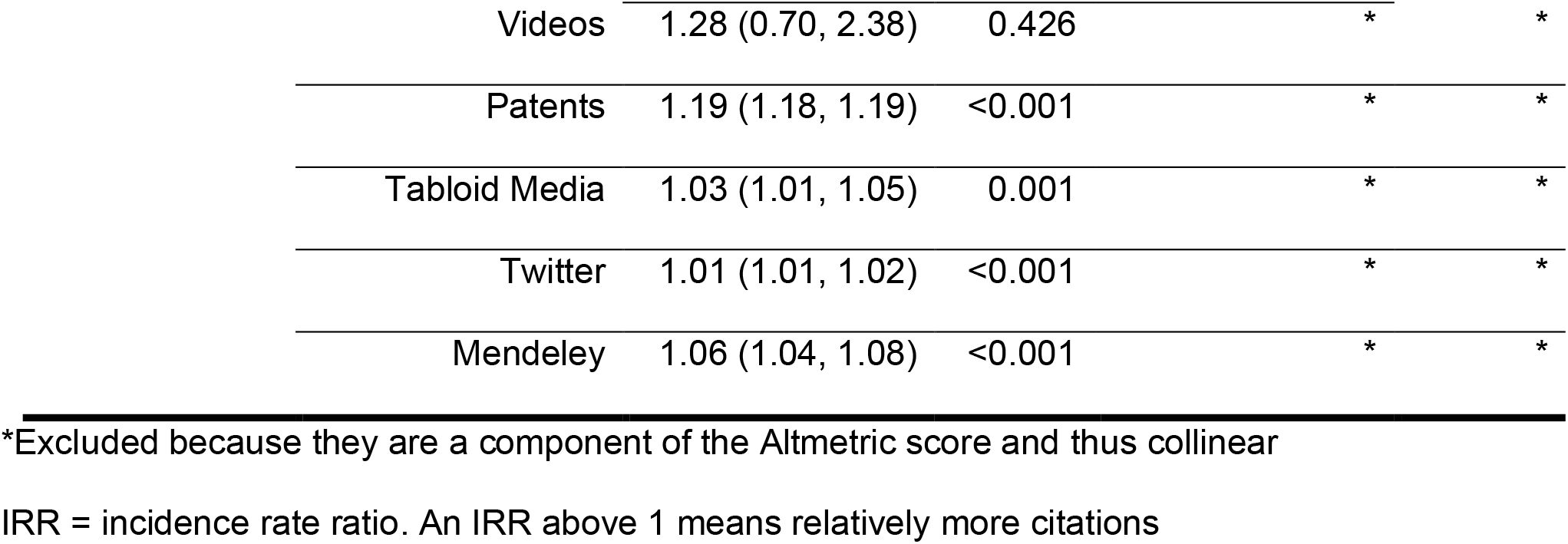
The relationship between citations, article characteristics and social media attention

**Figure 1.**
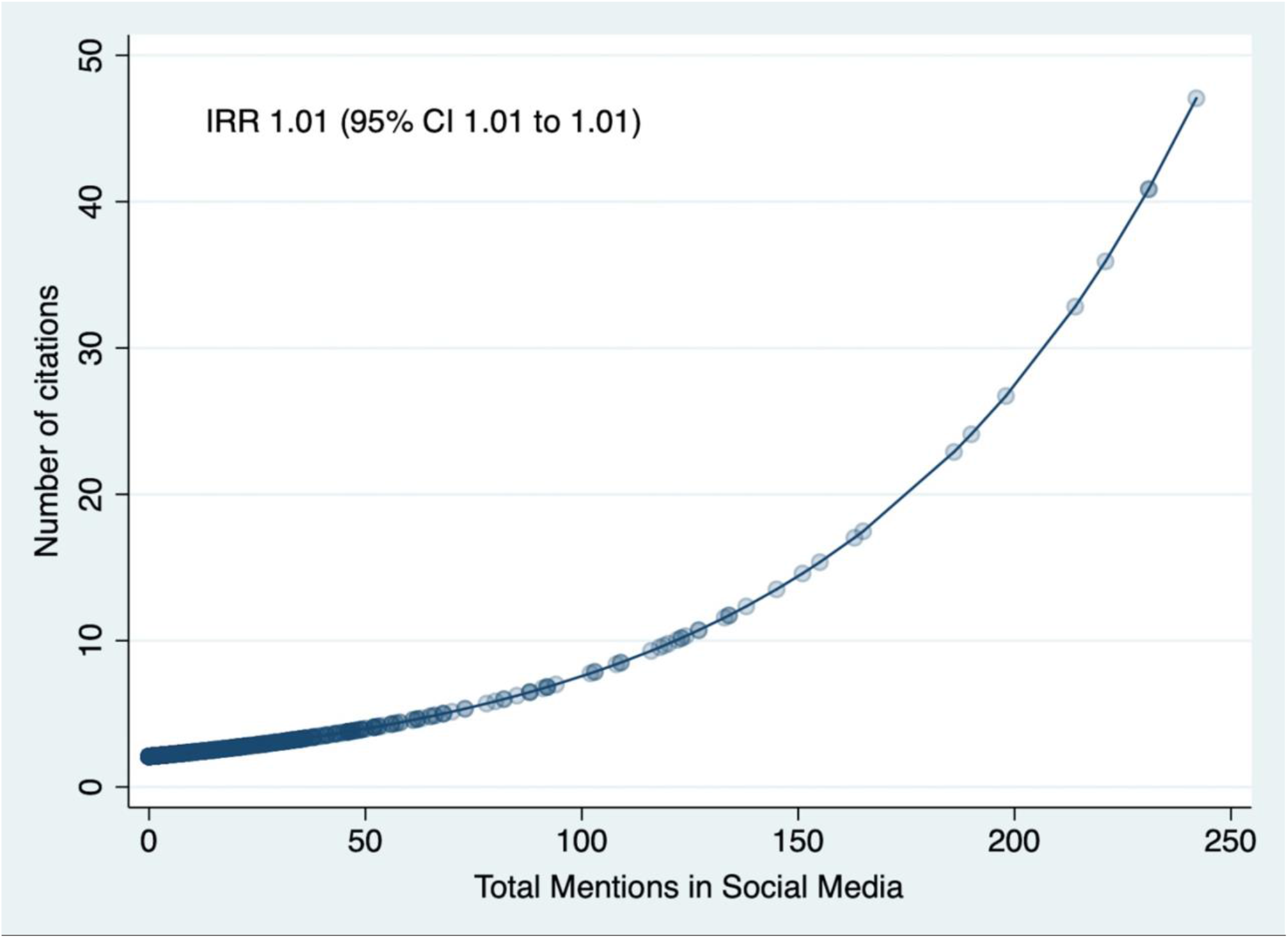
The relationship between the total number of mentions in social media and the citation rate of all articles.

**Figure 2.**
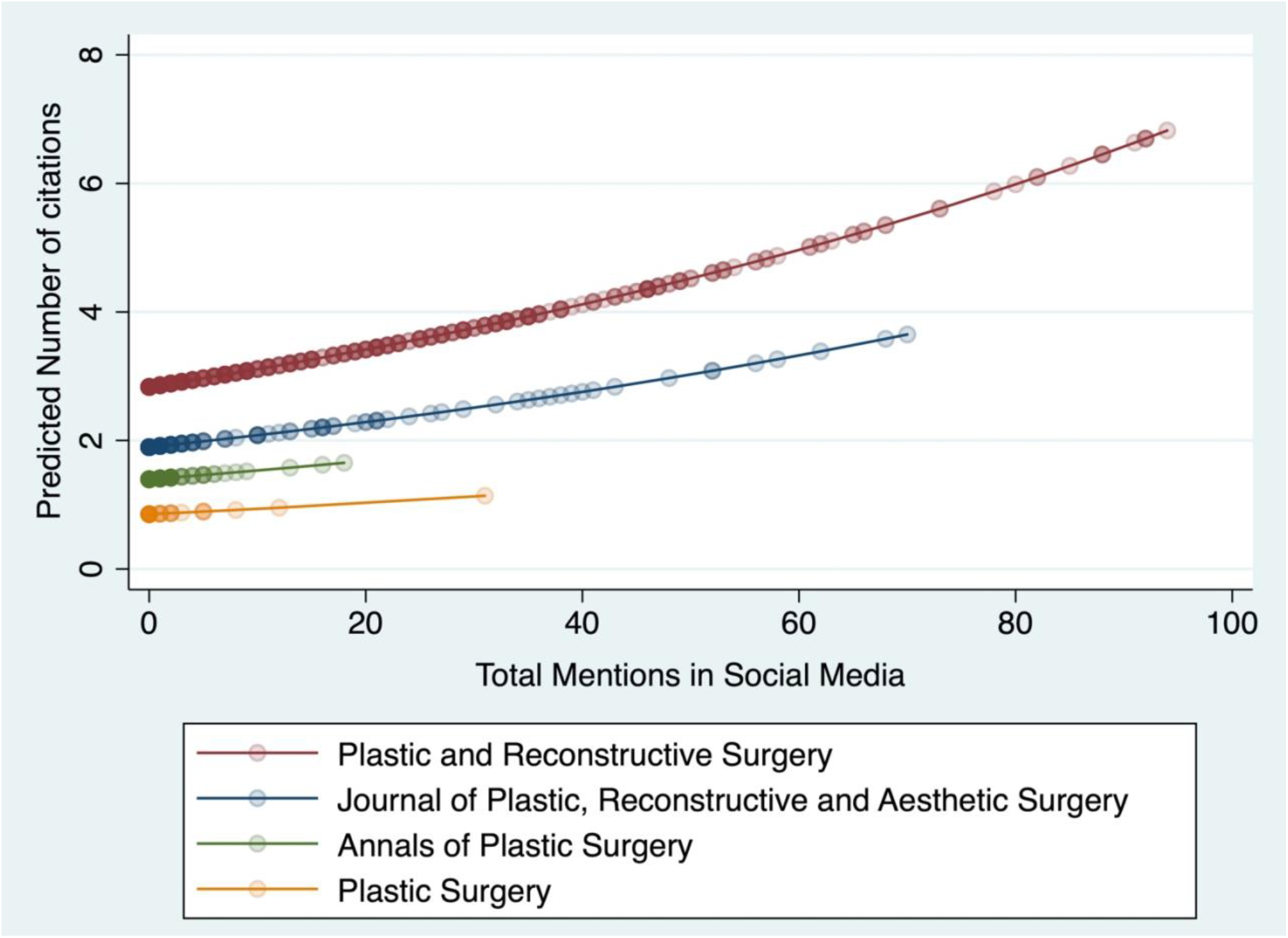
The relationship between the total number of mentions in social media and the citation rate of articles published in different journals.

**Figure 3.**
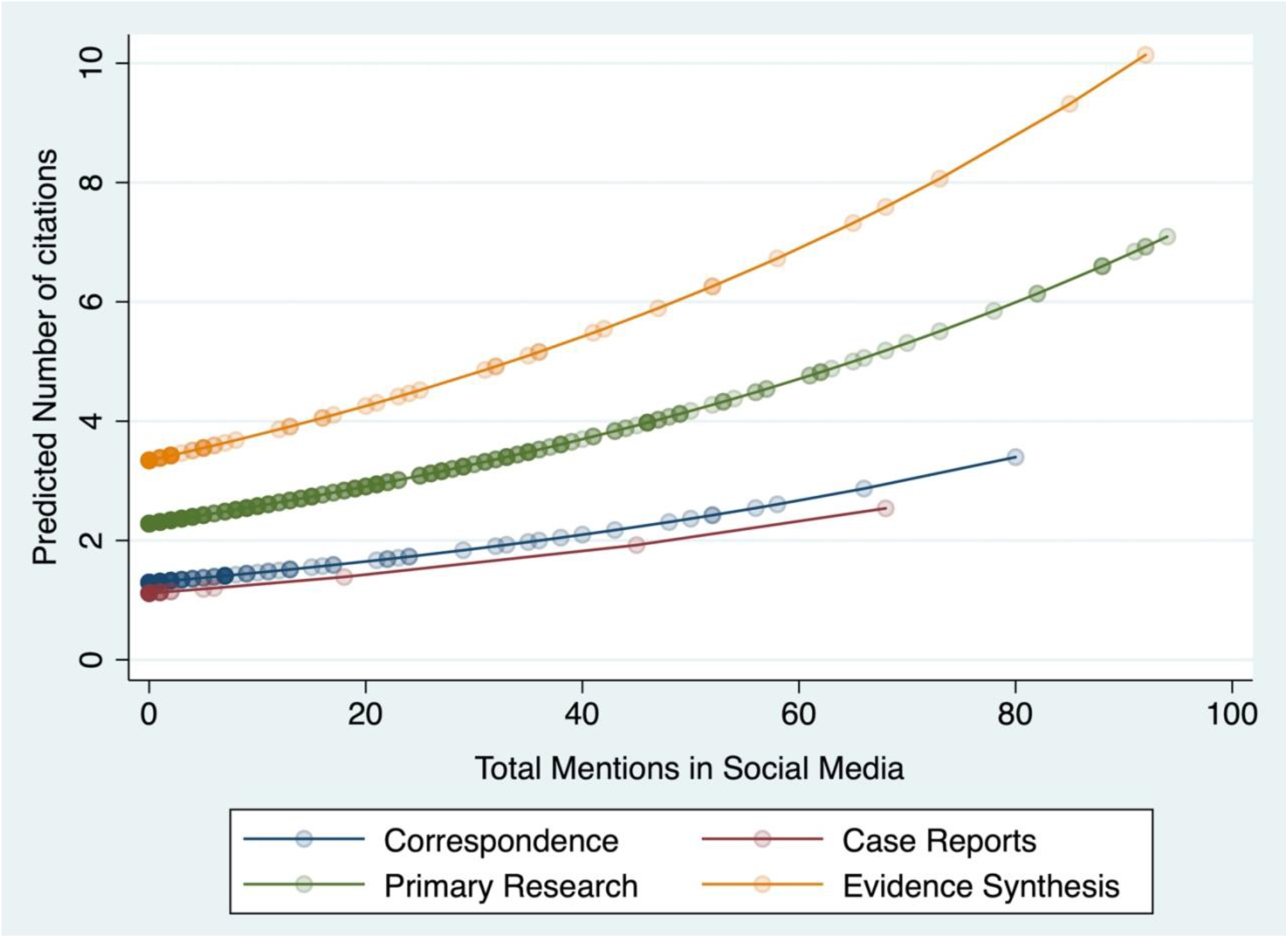
The relationship between the total number of mentions in social media and the citation rate of different types of articles.

## Discussion

We have shown that social media plays an important role in disseminating plastic surgery research. We present convincing evidence that plastic surgeons (and especially academic clinicians) and publishers should engage with social media to both maximise the impact of their research, and to filter out emergent and relevant articles from the ever-growing body of evidence.

This study adds to the literature concerning the interplay of SoMe and journals’ Impact Factor but re-opens the debate^5,9,10^ of *‘who came first?’* – do people follow and subscribe to journals on SoMe platforms because their articles are regularly cited, or do articles get cited more often because they are more widely seen and distributed on SoMe^9,11^? The direction and magnitude of the relationship between social impact and traditional metrics (citations) is difficult to elucidate. Several journals in the fields of trauma and orthopaedic surgery, urology, and radiology have shown a strong association between altmetrics and their respective Impact Factors^9,10,12^. Further, the Impact Factor of journals with a Twitter profile have consistently exhibited greater mean growth compared to journals without Twitter accounts^5,10,12^. Whilst recent work suggested no relationship between plastic surgery journals’ Impact Factor and their Twitter presence5, no evidence of an association is not the same as evidence of no association. Ultimately, given that Impact Factor is a function of citations over time, and our work shows that SoMe activity increases citation rates, we concur with the wider scientific community in stating that SoMe is a powerful force in the shaping author-, article- and journal-level metrics in plastic surgery.

Numerous articles over the last decade have shown that Tweets are a significant predictor of citations in non-specialty medical journals^13^. This is reflected in subspecialty journals also, where altmetrics are associated with inflated citation rates in the fields of cardiology^14^, emergency medicine^15^, gastroenterology^16^, general surgery^17,18^, paediatric surgery^19^, pain medicine^20^ and urology^7^. This is particularly true for humorous or curious topics^7,21,22^. Additionally, authors who promote their own research on SoMe receive more citations than those who do not^7^, which stands to reason given that SoMe is an advertising platform. Eysenbach showed that Tweets have a half-life and (what will be the) top-cited articles of the future can be predicted from the first 3-days of Twitter activity with 93% specificity and 75% sensitivity^13^. Academics should note that citations increase by 1% per 1.09% increase in Twitter followers, and Editors should note that journals recruiting a 1.46% increase in Twitter followers increase their Impact Factor by to 1%. Regardless of the impact on citation rates, SoMe is a powerful tool for discourse between academics, the medical community and the public. By circulating research findings via SoMe an author is able to widen the reach of their paper beyond the confines of a journal. We suggest that future researchers poll SoMe users interested in academic plastic surgery to create a new hashtag representing high-quality plastic surgery research.

Two other factors in our study appeared to affect research impact – the type of article and whether it is free to access (open access). Systematic reviews attracted the highest attention on SoMe and ultimately, citations too (Table 3 and Figure 3) which has been widely demonstrated in other fields^23^. This should serve as endorsement for authors and Editors alike, in the production and publication of high-quality and relevant systematic reviews in plastic surgery. it is widely accepted that freely available articles (published gold or green open-access) are read and cited more often^24,25^, experiencing up to an 18% uplift in citations^13,26^; however, in this study open-access publication was not associated with an increased rate of citations^21^. The readership of plastic surgery research is relatively small and largely limited to health professionals, many of whom will have access to articles published within the included journals via institutional or collegiate subscriptions, which may explain why we observed no association between open-access publication status and citations.

### Limitations

There are several competing companies which track and transform altmetrics including: Plum Analytics (PlumX), Altmetrics, ImpactStory and more. Data produced by Altmetric are more accurate long-term whilst PlumX metrics are more accurate when articles have zero-reads or one-read-only, as measured by Mendeley^26,27^. We decided that long-term count accuracy was most important so elected to use Altmetrics, although researchers should consider extracting data from both Altmetric and PlumX platforms for comparison.

The Impact Factor, SCImago Journal Rank and h-index are prone to bias^28^ because publications in the English language are more highly cited^28^ and citations take years to stabilise^29^; such biases may also exist for altmetrics which could weaken our inferences.

The pseudo-R2 of our multivariable negative binomial model was only 6% which suggests that there were other factors which influence whether or not articles were cited. The ‘appeal’ of an article, the quality of its construct, the reputation of the author(s) and their institution(s), whether it addresses a topical and important issue in the present time, etc., are difficult concepts to measure and so, impossible to model statistically, which limited the translation of our findings.

## Conclusions

We have shown that social media is an important part of disseminating plastic surgery research.

## Data Availability

The authours confirm that all relevant data are contained in the article

## Acknowledgements

None.

## Conflicts of Interest

There are no conflicts of interest.

## Funding

Ryckie Wade is a Doctoral Research Fellow funded by the National Institute for Health Research (NIHR, DRF-2018-11-ST2-028). The views expressed are those of the author(s) and not necessarily those of the NHS, the NIHR or the Department of Health and Social Care.

## Ethical review

Not applicable

## Disclosures

None

---

**Supplementary Figure 1.**
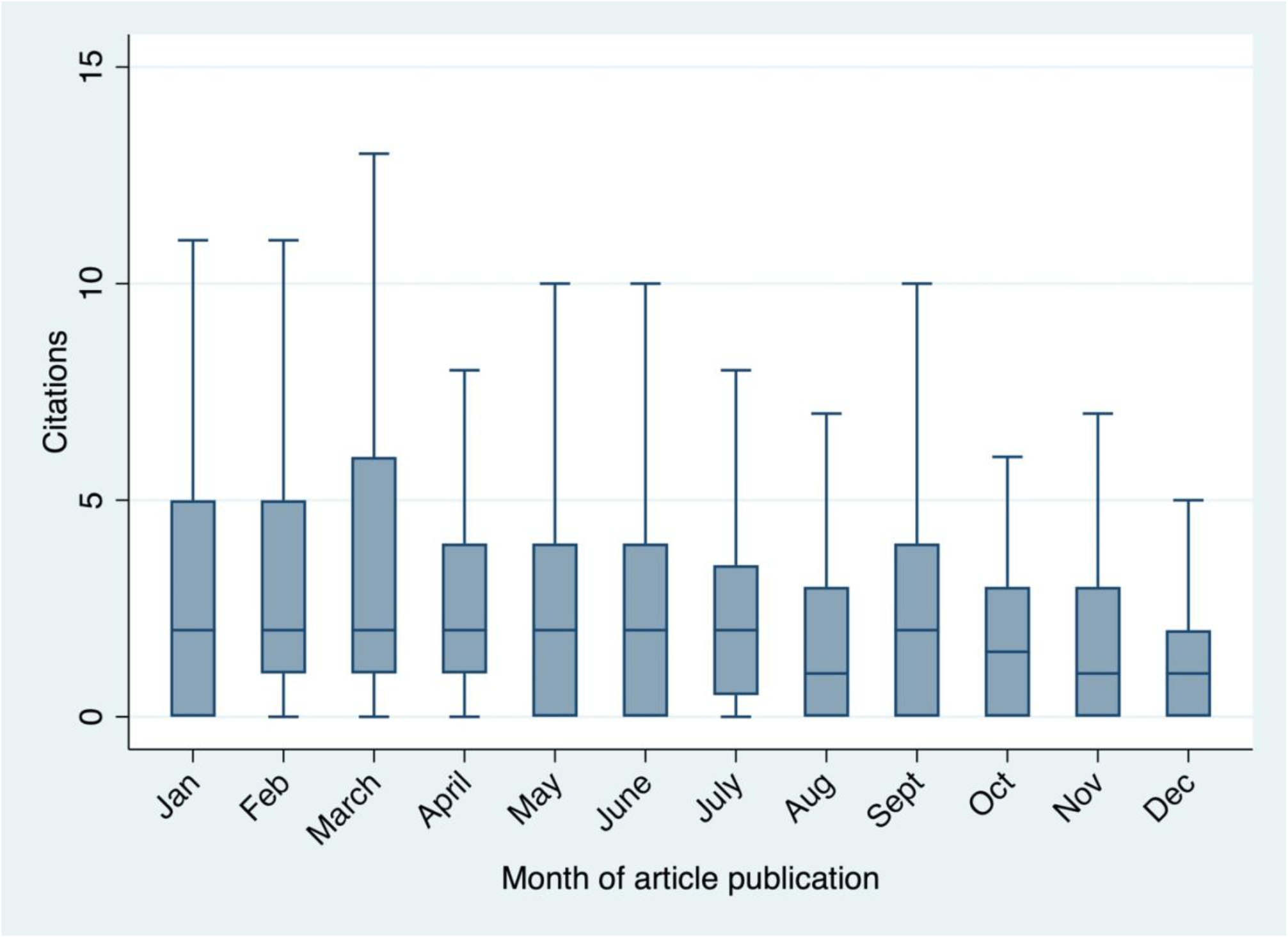
Boxplot of citation count according to the month in which articles were published. Articles published during August, November and December had significant fewer citations than other months of the year.

**Supplementary Table 1.** Weightings for the Altmetric Socre

| Source | Weighting |
| --- | --- |
| News | 8 |
| Blog | 5 |
| Policy document (per source) | 3 |
| Patent | 3 |
| Wikipedia | 3 |
| Twitter (tweets and retweets) | 1 |
| Peer review (Publons, Pubpeer) | 1 |
| Weibo (not trackable since 2015, but historical data kept) | 1 |
| Google+ (not trackable since 2019, but historical data kept) | 1 |
| F1000 | 1 |
| Syllabi (Open Syllabus) | 1 |
| LinkedIn (not trackable since 2014, but historical data kept) | 0.5 |
| Facebook (only a curated list of public Pages) | 0.25 |
| Reddit | 0.25 |
| Pinterest (not trackable since 2013, but historical data kept) | 0.25 |
| Q&A (Stack Overflow) | 0.25 |
| Youtube | 0.25 |
| Number of Mendeley readers | 0 |
| Number of Dimensions and Web of Science citations | 0 |

## Notes

### Competing Interest Statement

The authors have declared no competing interest.

